# Changes in hospital staff’ mental health during the Covid-19 pandemic: longitudinal results from the international COPE-CORONA study

**DOI:** 10.1101/2023.04.20.23288855

**Authors:** Roberta Lanzara, Chiara Conti, Ilenia Rosa, Tomasz Pawłowski, Monika Malecka, Joanna Rymaszewska, Piero Porcelli, Barbara Stein, Christiane Waller, Markus M. Müller, the Cope-Corona Study Group

**Affiliations:** Department of Psychological, Health, and Territorial Sciences, University “G. d’Annunzio” of Chieti-Pescara, Chieti, Italy; Department of Psychiatry, Wroclaw Medical University, Wrocław, Poland; Department of Psychosomatic Medicine and Psychotherapy, Paracelsus Medical University, General Hospital Nuremberg, Germany

## Abstract

This longitudinal study aimed to explore anxiety and depressive symptoms, individual resources, and job demands in a multi-country sample of 612 healthcare workers (HCWs) during the COVID-19 pandemic. Two online surveys were distributed to HCWs in seven countries (Germany, Andorra, Ireland, Spain, Italy, Romania, Iran) during the first (May-October 2020, T1) and the second (February-April 2021, T2) phase of the pandemic, assessing sociodemographic characteristics, contact with COVID-19 patients, anxiety and depressive symptoms, self-compassion, sense of coherence, social support, risk perception, and health and safety at the workplace. HCWs reported a significant increase in depressive and anxiety symptoms. HCWs with high depressive or anxiety symptoms at T1 and T2 reported a history of mental illness and lower self-compassion and sense of coherence over time. Risk perception, self-compassion, sense of coherence, and social support were strong independent predictors of depressive and anxiety symptoms at T2, even after controlling for baseline depressive or anxiety symptoms and sociodemographic variables. These findings pointed out that HCWs during the COVID-19 outbreak experienced a high burden of psychological distress. The mental health and resilience of HCWs should be supported during disease outbreaks by instituting workplace interventions for psychological support.

## Introduction

The outbreak of COVID-19 pandemic has become a major worldwide health crisis and has had a huge impact on almost every area of people’s life. In addition to the physical health consequences, public health experts from all over the world have raised concerns over the global mental health crisis due to quarantine, social isolation, loss, economic hardship, the threat of becoming infected, stigmatization, and uncertainty about the future [1,2]. These external factors resulted in people’s expanded vulnerability to manifest onset of symptoms and exacerbation of already existing psychopathology [3]. This situation came as no surprise in the context of previous epidemics, when numerous studies reported raised levels of psychological distress in the population undergoing such a crisis [4,5], leading to maladaptive behaviors [6–8]. A systematic review [9] assessed the pandemic’s psychological impact on the general population and affirmed that well-being has been lower with higher levels of depression, generalized anxiety symptoms, sleeping problems and distress compared to baseline measures. This systematic review found an association between higher levels of psychological distress and female gender, younger age, chronic medical or psychiatric illness, student status, unemployment, and constant exposure to news or social media. In addition, knowing someone with a COVID-19 infection, being at huge risk of contamination, outside-of-the-home work, or being under an obligatory stay-at-home order have been recognized as factors related to worse mental health, as has also been shown by other studies not included in the review [10–12].

To effectively play their role during the COVID-19 pandemic, it was essential for healthcare workers (HCWs) to maintain their psychological and mental well-being [13,14], as they were standing at the frontline of the crisis. However, the evidence has shown that the wide spread of COVID-19 has had a remarkable impact on their mental health (such as stress or burnout) [13,15–18]. Anxiety, insomnia, and somatic complaints were more common among HCWs than among non-medical workers, according to certain meta-analyses and systematic reviews [19–22]. Results from other studies have shown that at least one in five HCWs reported symptoms of depression and anxiety, and almost 4/10 HCWs reported sleep problems [23].

So far, researchers have noticed many stress-related responses among the population of HCWs that were both adaptive and maladaptive [24]. These responses can affect one’s psychological comfort and can turn out to be either protective or risk factors for unfavorable mental-health outcomes. What is worth mentioning, there were reported higher scores for positive coping strategies than negative ones among HCWs, indicating that health care personnel are able to employ positive coping strategies when faced with stressful situations like the COVID-19 pandemic. Some studies have explored individual differences in psychological risk and protective factors and their connections with COVID-19-related mental health among HCWs. Utilization of healthy coping strategies such as asking for social support, positive thinking and problem solving was linked to lower levels of traumatic stress, stigma, psychological distress, stress symptoms, anxiety, and depression, as well as psychological distress [24].

Psychological resilience – a construct representing the ability to cope with set-backs and adapt positively in the face of adversity [25] – was connected to better mental well-being during the pandemic and less burnout among HCWs, especially nurses. It is also worth noting that as many as about half of them showed symptoms of moderate or severe burnout. Additionally, resilience scores showed statistically significant inverse relationships with PTSD, anxiety, and depression [26].

Some meta-analyses have shown that self-compassion was negatively associated with psychopathology and positively with well-being [27–28]. The study conducted by Hi-Po Lau et al. [30] confirmed the previous findings: self-compassion buffered the association between perceived COVID-related threats and psychological distress. For risk factors associated with feeling higher levels of mental-health burden during COVID-19 pandemic, the following were listed: disgust sensitivity, anxiety sensitivity, body vigilance, concerns of being infected and general distress [31].

Finally, several nonadaptive personality traits including negative affectivity and detachment were related to higher levels of depression, anxiety, and stress during the COVID-19 pandemic [10,32].

Accordingly, our study aimed to investigate long-term changes in levels of distress and protective and risk factors in hospital staff during the COVID-19 pandemic. Specifically, our aim was twofold: (1) to assess the sociodemographic factors, job demands (contact with COVID-19 patients, risk perception, health and safety on the workplace), and individual resources (self-compassion, sense of coherence, social support) associated with changes in anxiety and depression symptoms during the COVID-19 pandemic; (2) to explore whether and to what extent sociodemographic factors, job demands, and individual resource predicted high levels of anxiety and depressive symptoms during the COVID-19 pandemic. Based on previous findings, it was hypothesised that: (1) HCWs with high levels of anxiety and depression over time would show higher levels of job demands and lower individual resources during both the first and second phases of the COVID-19 pandemic; (2) higher job demands and lower individual resources would predict higher levels of depression and anxiety during the second phase of the COVID-19 pandemic.

## Materials and Methods

### Procedure and participants

This study is part of the larger Cope-Corona project aimed at assessing protective and risk factors for medical staff during the COVID-19 pandemic [for more details see (17)]. The project was conducted with the support of the European Association of Psychosomatic Medicine (EAPM), along with the coordination of Paracelsus Medical University, Nuremberg General Hospital, and Catholic University Eichstätt-Ingolstadt. All EAPM members were informed of the research initiative and were invited to participate. As a result of this process, an international research group was established. Data presented in this study were provided by seven centers: Hospital Nostra Senyora de Meritxell (Andorra), General Hospital Nuremberg (Germany), Tehran University of Medical Sciences (Iran), Private Health Sector and Public Health Sector (Ireland), University G. d’Annunzio of Chieti and Pescara (Italy), Dexeus University Hospital of Barcelona and Hospital Clínic de Barcelona (Spain), University of Medicine and Pharmacy Iuliu Hatieganu, Cluj-Napoca (Romania). Additional data from China and Poland were available cross-sectionally and were not included in the current study based on longitudinal data.

Two online surveys were distributed to hospital staff during the first (May-October 2020, T1) and the second (February-April 2021, T2) phase of the pandemic. The participants were invited through email to complete the online survey (www.qualtrics.com/it) on both occasions. To optimize ecological validity, all adult (years≥18) employees of the hospitals and their subcontractors were included. To ensure the validity of the responses, the inclusion criterion was a response of at least 50% of the questions. After removing those who did not satisfy the inclusion criteria, a total of 2,097 and 4,240 participants were enrolled at T1 and T2, respectively. By matching the self-generated code, a total of 612 participants were identified for longitudinal analysis.

### Measures

Study variables were measured using validated psychometric scales or with ad hoc instruments where adequate measures were not available. All scales were set to local languages (Catalan, English, Farsi, German, Italian, and Spanish) using validated translations when available. When translations were not available, items were translated and back-translated into the original language. Factor and reliability analyses were used to test all ad hoc scales [for details, see(17)].

### Sociodemographic characteristics

Ad hoc questions regarding sociodemographic and occupational variables were included in the online survey. Data were self-reported by participants, including age, gender, work experience, and function (physicians, nurses, technicians, administrative, others). History of mental illness was assessed with a single item by asking whether participants had ever suffered from a mental disorder at any time in their life (“yes” or “no”).

### Job demands

#### Contact with COVID-19 patients

Professional contact with COVID-19 patients was assessed with a single item by asking whether participants were directly involved in the clinical management of patients with suspected or confirmed coronavirus infection. The answer rated on a 4-point Likert-scale ranging from “not at all” to “very much”. Contact with COVID-19 patients and possible changes between T1 to T2 were coded as “any”, “increase in contact”, “decrease in contact”, “much”. The dichotomous variable of contact with COVID-19 patients was used in the comparisons, coded as no (never had contact at both T1 and T2) and yes (contact at T1 and/or T2).

#### Risk perception

The risk perception COVID-19-related was assessed with three items regarding the probability of becoming infected, concern about becoming severely infected, and concern about infecting others. Each item was rated on a 5-point Likert-scale. The total score was calculated by the mean of three items, with higher scores indicating higher levels of risk perception.

#### Health and safety on the workplace

The subjective feelings about health and safety at the workplace were assessed with two items regarding the availability of personal protective equipment, and confidence on stay healthy at work. Both items were rated on a 5-point Likert-scale. The total score was calculated by the mean of two items reversed, with higher scores indicating higher feelings of health and safety.

### Individual resources

#### Self-Compassion

The State Self-Compassion Scale-Short form (SSCS-S) was used to assess the tendency to respond in a self-compassionate way to a current painful situation or life difficulty [34]. The SCSS-S is a 6-item questionnaire rated on 5-point Likert-scale ranging from

1 (“not at all true for me”) to 5 (“pretty much true for me”). The total score was calculated by the sum of the six items, with higher scores indicating higher levels of self-compassion.

#### Sense of Coherence

The 3-item Sense of Coherence (SOC-3) questionnaire was used to assess the tendency to have an enduring and dynamic sense of confidence based on three components: understandability, manageability, and meaning [35,36]. The SOC is rated on 7- point Likert-scale. The total score was calculated by the sum of three items, with higher scores indicating higher levels of sense of coherence.

#### Social support

The ENRICHD Social Support Inventory (ESSI) is a 5-item scale used to evaluate four domains of social support: emotional, instrumental, informational, and appraisal [37]. Respondents are asked to estimate how helpful they perceive people close to them to be on a 5-point Likert-scale ranging from 1 (“none of the time”) to 5 (“all of the time”). The total score was calculated by the sum of five items. A score of ≤3 on at least 2 of the 5 items with a total score of ≤18 indicates a lack of social support.

### Emotional distress

#### Depressive symptoms

The 2-item Patient Health Questionnaire (PHQ-2) was used to assess the presence of depression symptoms [38]. The PHQ-2 is a self-report measure designed to screen for depressed mood and anhedonia over the past two-weeks. Each item is rated on a 4- point Likert-scale, with total scores varying from 0 (“not at all”) to 3 (“nearly every day”). The total score was calculated by the sum of two items. A score of 3 or above is used as the cutoff for greater severity of depression. To evaluate the possible changes in depressive symptoms between T1 to T2, the PHQ-2 scores were coded as “never” (no symptoms at both T1 and T2), “decrease in symptoms” (high symptoms at T1), “increase in symptoms” (high symptoms at T2), “always” (high symptoms at both T1 and T2).

#### Anxiety symptoms

The 2-item General Anxiety Disorder-2 (GAD-2) was used to measure the presence of anxiety symptoms [39]. The GAD-2 is a self-report questionnaire used to assess feeling nervous and not being able to control thoughts/worries, two core criteria for anxiety.

Respondents are asked to estimate the frequency of these symptoms over the past two-weeks on a 4-point Likert-scale ranging from 0 (“not at all”) to 3 (“nearly every day”). The total score was calculated by the sum of two items. A score of 3 or above is used as the cutoff for greater severity of anxiety. To evaluate the possible changes in anxiety symptoms between T1 to T2, the GAD-2 scores were coded as “never” (no symptoms at both T1 and T2), “decrease in symptoms” (high symptoms at T1), “increase in symptoms” (high symptoms at T2), “always” (high symptoms at both T1 and T2).

### Statistical analyses

Data analysis was performed using SPSS 26.0. Descriptive statistics were reported in terms of frequencies.

A 3-step strategy was used for data analysis.

First, a paired-sample Student’s t-test was used to evaluate within-subjects differences in emotional distress in the two time-points assessments. The standardized mean difference (Cohen’s d) was used as a measure of effect size considered as small (0.20–0.50), moderate (0.50–0.80), and large (>0.80) [40].

Second, two chi-square (χ^2^) tests and two repeated-measures analyses of variance (ANOVAs) were used to compare between-group differences in sociodemographic characteristics, job demands, and individual resources for depression-related groups and anxiety-related groups during the two time-points. The repeated-measures ANOVAs included measures of anxiety and depressive symptoms, self-compassion, sense of coherence, social support, risk perception, and health and safety as the dependent variable, the time points T1 and T2 as a within-subject factor, and depression-related/anxiety-related groups as the between-subject comparison. Effect sizes for categorical variables were assessed by using Cramér’s V ranging from 0 to +1, where 0 indicates the complete independence of two variables and +1 indicates a perfect association. The Eta-squared (η^2^) was used as a measure of effect size for continuous variables. A standardized effect size of 0.01–0.05 is considered small, 0.06–0.14 moderate, and >0.14 large [41].

Third, two hierarchical regressions were used to identify those factors that best predict depressive symptoms and anxiety symptoms at T2. The PHQ-2 and GAD-2 scores at T2 were considered as the dependent variable and the independent variables were the baseline GAD-2 or PHQ-2, age, gender, function, job experience, history of mental illness, contact with COVID-19 patients, risk perception (T1,T2), health and safety (T1,T2), SCS (T1,T2), SOC (T1,T2), and ESSI (T1,T2). The predictors were entered in separate blocks to determine how well each variable predicted the outcome.

## Results

### Characteristics of the sample

The participants were mostly women (n=462, 75.5%), nurses (n=182, 29.7%), with more than 35 years (n=446, 72.9%) and 6 years of job experience (n=474, 77.6%), and had not a history of mental illness (n=474, 77.4%) or contact with COVID-19 patients at neither T1 nor T2 (n=358, 58.6%) (see Table1). The number of participants for each country was reported in Table S1.

Participants reported a significant worsening of depressive (d=0.21) and anxiety (d=0.25) symptoms during the two phases of pandemic-related restrictions (see Table S2).

Overall, 59.9% and 71.2% of the sample reported anxiety or depressive symptoms during the two waves of the COVID-19 pandemic (see Table 1). Specifically: 1) 32.6% and 39.8% of the sample reported symptoms of anxiety or depressive symptoms during the first and second waves of the pandemic; 2) 17.8% and 21.9% reported an increase in symptoms during the second wave; 3) and 9.5% reported a decrease in symptoms during the second wave.

**Table 1.** Comparisons of sociodemographic and occupational characteristics in total sample, depression-related and anxiety-related groups.

| Variable, N (%) | Total sample<br>N = 612 | Depression-related groups<sup>a</sup> | | | | $\chi^2$ | Cramer's<br>V | Anxiety-related groups<sup>a</sup> | | | | $\chi^2$ | Cramer's<br>V |
| --- | --- | --- | --- | --- | --- | --- | --- | --- | --- | --- | --- | --- | --- |
|  |  | Never<br>N = 177<br>(28.8%) | Decreased<br>N = 58<br>(9.5%) | Increased<br>N = 134<br>(21.9%) | Always<br>N = 243<br>(39.8%) |  |  | Never<br>N = 246<br>(40.1%) | Decreased<br>N = 58<br>(9.5%) | Increased<br>N = 109<br>(17.8%) | Always<br>N = 199<br>(32.6%) |  |  |
| Age |  |  |  |  |  |  |  |  |  |  |  |  |  |
| ≤35 years | 166 (27.1) | 38 (21.5) | 15 (25.9) | 42 (31.3) | 71 (29.2) | 4.66 | .09 | 62 (25.2) | 13 (22.4) | 29 (26.6) | 62 (31.2) | 2.76 | .07 |
| >35 years | 446 (72.9) | 139 (78.5) | 43 (74.1) | 92 (68.7) | 172 (70.8) |  |  | 183 (74.8) | 45 (77.6) | 80 (73.4) | 137 (68.8) |  |  |
| Gender |  |  |  |  |  |  |  |  |  |  |  |  |  |
| Men | 150 (24.5) | 53 (29.9) | 12 (20.7) | 28 (20.9) | 57 (23.5) | 4.37 | .08 | 74 (30.1) | 12 (20.7) | 26 (23.9) | 38 (19.1) | 6.53 | .11 |
| Women | 462 (75.5) | 124 (70.1) | 46 (79.3) | 106 (79.1) | 186 (76.5) |  |  | 172 (69.9) | 46 (79.3) | 83 (76.1) | 161 (80.9) |  |  |
| Function |  |  |  |  |  |  |  |  |  |  |  |  |  |
| Physicians | 84 (13.7) | 30 (16.9) | 6 (10.3) | 20 (14.9) | 28 (11.5) | 11.91 | .08 | 42 (17.1) | 10 (17.2) | 16 (14.7) | 16 (8) | 22.77* | .11 |
| Nurses | 182 (29.7) | 45 (25.4) | 17 (29.3) | 46 (34.3) | 74 (30.5) |  |  | 72 (29.3) | 20 (34.6) | 34 (31.2) | 56 (28.1) |  |  |
| Technicians | 79 (12.9) | 24 (13.6) | 6 (10.3) | 12 (9) | 37 (15.2) |  |  | 31 (12.6) | 9 (15.5) | 12 (11) | 27 (13.6) |  |  |
| Administrative | 147 (24) | 48 (27.2) | 15 (25.9) | 33 (24.6) | 51 (21) |  |  | 64 (26) | 9 (15.5) | 30 (27.5) | 44 (22.2) |  |  |
| Others<sup>1</sup> | 120 (19.6) | 30 (16.9) | 14 (24.1) | 23 (17.2) | 53 (21.8) |  |  | 37 (15) | 10 (17.2) | 17 (15.6) | 56 (28.1) |  |  |
| Job experience |  |  |  |  |  |  |  |  |  |  |  |  |  |
| ≤6 years | 137 (22.4) | 35 (19.8) | 11 (19) | 26 (19.4) | 65 (26.7) | 4.43 | .08 | 45 (18.3) | 9 (15.5) | 22 (20.2) | 61 (30.6) | 12.08** | .14 |
| >6 years | 475 (77.6) | 142 (80.2) | 47 (81) | 108 (80.6) | 178 (73.3) |  |  | 201 (81.7) | 49 (84.5) | 87 (79.8) | 138 (69.4) |  |  |
| Contact with<br>COVID-19<br>patients |  |  |  |  |  |  |  |  |  |  |  |  |  |
| No | 358 (58.5) | 108 (61) | 38 (65.5) | 78 (58.2) | 134 (55.1) | 2.77 | .07 | 154 (62.6) | 32 (55.2) | 63 (57.8) | 109 (54.8) | 3.13 | .07 |
| Yes | 254 (41.5) | 69 (39) | 20 (34.5) | 56 (41.8) | 109 (44.9) |  |  | 92 (37.4) | 26 (44.8) | 46 (42.2) | 90 (45.2) |  |  |
| History of mental<br>illness |  |  |  |  |  |  |  |  |  |  |  |  |  |
| Yes | 138 (22.6) | 20 (11.9) | 10 (17.2) | 27 (20.1) | 81 (33.3) | 30.38*** | .22 | 74 (30.1) | 12 (20.7) | 26 (23.8) | 38 (19.1) | 39.42*** | .25 |
| No | 474 (77.4) | 157 (88.1) | 48 (82.8) | 107 (79.9) | 162 (66.7) |  |  | 172 (69.9) | 46 (79.3) | 83 (76.2) | 161 (80.9) |  |  |

### Between-group comparisons

The results of comparisons of sociodemographic and occupational factors in depression-related and anxiety-related groups are reported in Table 1.

Only the history of mental illness (χ^2^=30.38, p<.001) showed significant between depression-related groups differences, with the effect size in the small range.

There were significant differences between anxiety-related groups on gender (χ^2^=6.53, p=.09), function (χ^2^=22.77, p=.03), job experience (χ^2^=12.08, p=.007), and history of mental illness (χ^2^=39.42, p<.001), with effect sizes in the small ranges.

The results of repeated measure ANOVA in depression-related groups are reported in Table 2.

**Table 2.** Comparisons of anxiety symptoms, job demands, and individual resources over time in depression-related groups.

| Variables,<br>mean (SD) | Depression-related groups<sup>a</sup> |  |  |  | Time |  | Time*Group |  | Within-Groups<br>comparisons |
| --- | --- | --- | --- | --- | --- | --- | --- | --- | --- |
| | A<br>Never<br>N = 177 | B<br>Decreased<br>N = 58 | C<br>Increased<br>N = 134 | D<br>Always<br>N = 243 | F | $\eta^2$ | F | $\eta^2$ | Post-hoc<br>Bonferroni |
| GAD-2 |  |  |  |  |  |  |  |  |  |
| T1 | 0.73 (0.90) | 1.56 (1.18) | 0.87 (0.97) | 2.32 (1.46) | 0.59 | .001 | 22.39*** | .11 | A < B, C, D;<br>D > A, B, C |
| T2 | 0.72 (0.83) | 0.87 (0.85) | 1.71 (1.19) | 2.36 (1.39) |  |  |  |  |  |
| SSCS-S |  |  |  |  |  |  |  |  |  |
| T1 | 22.68 (3.01) | 20.67 (2.72) | 21.64 (3.59) | 19.38 (3.66) | 0.09 | - | 3.99** | .02 | A > B, C, D;<br>D < A, B, C |
| T2 | 22.55 (3.21) | 21.67 (3.29) | 20.89 (3.51) | 19.09 (3.50) |  |  |  |  |  |
| SOC-3 |  |  |  |  |  |  |  |  |  |
| T1 | 13.85 (2.14) | 12.74 (2.90) | 13.30 (2.52) | 12.08 (3.08) | 0.11 | - | 3.12* | .02 | A > C, D;<br>D < A, B, C |
| T2 | 13.86 (2.05) | 13.48 (2.48) | 12.72 (2.65) | 12.07 (2.91) |  |  |  |  |  |
| ESSI |  |  |  |  |  |  |  |  |  |
| T1 | 22.38 (2.77) | 20.18 (3.89) | 21.42 (3.81) | 19.61 (4.50) | 0.85 | .001 | 2.17 | .01 | A > B, C, D;<br>D < A, C |
| T2 | 22.37 (2.83) | 20.61 (3.99) | 20.79 (3.77) | 19.31 (4.61) |  |  |  |  |  |
| Risk<br>perception |  |  |  |  |  |  |  |  |  |
| T1 | 3.05 (0.72) | 3.09 (0.75) | 3.17 (0.71) | 3.32 (0.69) | 12.95*** | .02 | 1.31 | .01 | A < D |
| T2 | 3.09 (0.74) | 3.22 (0.70) | 3.29 (0.80) | 3.49 (0.72) |  |  |  |  |  |
| Health and<br>safety |  |  |  |  |  |  |  |  |  |
| T1 | 4.13 (0.69) | 4.04 (0.80) | 4 (0.73) | 3.67 (0.87) | 49.56*** | .08 | 0.61 | .003 | A > D;<br>D < A, B, C |
| T2 | 4.34 (0.56) | 4.27 (0.68) | 4.22 (0.60) | 3.97 (0.72) |  |  |  |  |  |

<sup>a</sup>Never = absence of anxiety symptoms at both T1 and T2; Decreased = presence of anxiety symptoms at T1 but no longer at T2; Increased = presence of anxiety symptoms at T2 but not at T1; Always = presence of anxiety symptoms at both T1 and T2.
Notes: GAD-2 = General Anxiety Disorder-2; SSCS-S = Self-Compassion Scale; SOC-3 = Sense of Coherence; ESSI = ENRICH Social Support Instrument.
All values are standardized regression weights. \* $p < .05$ ; \*\* $p < .01$ ; \*\*\* $p < .001$ .

A significant effect of time on risk perception (F=12.95, p <.001) and health and safety (F=49.56, p<.001) was found. Comparing depression-related groups, participants with high depressive symptoms at both T1 and T2 (group D) reported higher anxiety symptoms (F=22.39, p<.001), lower self-compassion (F=3.99, p=.008), and lower sense of coherence over time (F=3.12, p=.03) than other groups.

The results of repeated measure ANOVA in anxiety-related groups are reported in Table 3.

**Table 3.** Comparisons of depressive symptoms, job demands, and individual resources in anxiety-related groups.

| Variables,<br>mean (SD) | Anxiety-related groups<sup>a</sup> |  |  |  | Time |  | Time*Group |  | Within-Groups<br>comparisons |
| --- | --- | --- | --- | --- | --- | --- | --- | --- | --- |
|  | A<br>Never<br>N = 246 | B<br>Decreased<br>N = 58 | C<br>Increased<br>N = 109 | D<br>Always<br>N = 199 | F | η<sup>2</sup> | F | η<sup>2</sup> | Post-hoc<br>Bonferroni |
| PHQ-2 |  |  |  |  |  |  |  |  |  |
| T1 | 0.89 (0.88) | 1.93 (1.33) | 1.17 (1.01) | 2.39 (1.33) | 7.46** | .01 | 26.86*** | .12 | A < B, C, D;<br>D > A, B, C |
| T2 | 1.12 (1.04) | 1.17 (1.04) | 2.23 (1.18) | 2.53 (1.31) |  |  |  |  |  |
| SSCS-S |  |  |  |  |  |  |  |  |  |
| T1 | 22.53 (3.11) | 20.81 (2.85) | 21.60 (2.89) | 18.74 (3.74) | 0.68 | .001 | 6.49*** | .03 | A > B, C, D;<br>D < A, B, C |
| T2 | 22.46 (3.14) | 21.83 (2.75) | 20.38 (3.22) | 18.51 (3.57) |  |  |  |  |  |
| SOC-3 |  |  |  |  |  |  |  |  |  |
| T1 | 13.99 (1.91) | 12.26 (3.06) | 13.31 (2.27) | 11.59 (3.25) | 1.33 | .002 | 9*** | .05 | A > B, C, D<br>D < A, B, C |
| T2 | 13.84 (2.06) | 13.79 (2.40) | 12.52 (2.76) | 11.59 (2.85) |  |  |  |  |  |
| ESSI |  |  |  |  |  |  |  |  |  |
| T1 | 21.76 (3.30) | 20.81 (4.55) | 21.13 (3.68) | 19.68 (4.54) | 2.42 | .004 | 2.49 | .01 | A > D |
| T2 | 21.61 (3.34) | 21.07 (4.62) | 20.27 (3.88) | 19.57 (4.61) |  |  |  |  |  |
| Risk<br>perception |  |  |  |  |  |  |  |  |  |
| T1 | 2.93 (0.68) | 3.34 (0.65) | 3.23 (0.67) | 3.43 (0.70) | 15.67*** | .03 | 1.11 | .01 | A < B, C, D |
| T2 | 2.99 (0.75) | 3.44 (0.70) | 3.43 (0.73) | 3.56 (0.67) |  |  |  |  |  |
| Health and<br>safety |  |  |  |  |  |  |  |  |  |
| T1 | 4.11 (0.75) | 3.81 (0.73) | 3.98 (0.77) | 3.64 (0.85) | 64.29*** | .10 | 2.11 | .01 | A > D<br>D < A, B, C |
| T2 | 4.35 (0.56) | 4.27 (0.59) | 4.13 (0.59) | 3.90 (0.75) |  |  |  |  |  |

<sup>a</sup>Never = absence of anxiety symptoms at both T1 and T2; Decreased = presence of anxiety symptoms at T1 but no longer at T2; Increased = presence of anxiety symptoms at T2 but not at T1; Always = presence of anxiety symptoms at both T1 and T2.
Notes: PHQ-2 = Patient Health Questionnaire-2; SSCS-S = Self-Compassion Scale; SOC-3 = Sense of Coherence; ESSI = ENRICHD Social Support Instrument.
All values are standardized regression weights. \*p < .05; \*\*p < .01; \*\*\*p < .001.

A significant effect of time on depression (F=7.46, p=.006), risk perception (F=15.67, p<.001) and health and safety (F=64.39, p<.001) was found. Comparing anxiety-related groups, participants with high anxiety symptoms at both T1 and T2 (group D) reported higher depressive symptoms (F = 26.86, p < .001), lower self-compassion (F=6.49, p<.001), and lower sense of coherence over time (F=9, p<.001) than other groups.

### Predicting depressive and anxiety symptoms at follow-up

A hierarchical regression analysis was performed to assess which variables contribute to explain the depressive symptoms at the T

2 (see Table 4).

**Table 4.** Summary of hierarchical regression model predicting depressive symptoms (PHQ-2) at T2.

| Variables | Model 1 |  |  | Model 2 |  |  | Model 3 |  |  | Model 4 |  |  |
| --- | --- | --- | --- | --- | --- | --- | --- | --- | --- | --- | --- | --- |
| | B | SE | $\beta$ | B | SE | $\beta$ | B | SE | $\beta$ | B | SE | $\beta$ |
| PHQ-2 (T1) | .50 | .04 | .49*** | .48 | .04 | .47*** | .44 | .04 | .43*** | .35 | .04 | .34*** |
| Age |  |  |  | -.15 | .14 | -.05 | -.11 | .14 | -.04 | -.03 | .13 | -.01 |
| Gender |  |  |  | -.01 | .11 | -.002 | -.02 | .11 | -.01 | -.07 | .10 | -.02 |
| Function |  |  |  | -.07 | .04 | -.07 | -.07 | .04 | -.03 | -.07 | .04 | -.07 |
| Job experience |  |  |  | -.13 | .16 | -.04 | -.07 | .16 | .02 | -.12 | .15 | -.04 |
| History of mental illness |  |  |  | .20 | .12 | .06 | .21 | .12 | .07 | .12 | .11 | .04 |
| Contact with COVID-19 patients |  |  |  |  |  |  | -.01 | .11 | -.003 | .03 | .10 | .01 |
| Risk perception (T1) |  |  |  |  |  |  | -.03 | .09 | -.02 | -.05 | .08 | -.03 |
| Risk perception (T2) |  |  |  |  |  |  | .24 | .08 | .14** | .20 | .08 | .11* |
| Health and safety (T1) |  |  |  |  |  |  | -.04 | .07 | -.03 | -.02 | .07 | -.01 |
| Health and safety (T2) |  |  |  |  |  |  | -.14 | .09 | -.07 | -.06 | .08 | -.03 |
| SSCS-S (T1) |  |  |  |  |  |  |  |  |  | .01 | .02 | .03 |
| SSCS-S (T2) |  |  |  |  |  |  |  |  |  | -.09 | .02 | -.24*** |
| SOC-3 (T1) |  |  |  |  |  |  |  |  |  | .02 | .02 | .04 |
| SOC-3 (T2) |  |  |  |  |  |  |  |  |  | -.09 | .02 | -.18*** |
| ESSI (T1) |  |  |  |  |  |  |  |  |  | .04 | .02 | .11* |
| ESSI (T2) |  |  |  |  |  |  |  |  |  | -.06 | .02 | -.19*** |
| **R<sup>2</sup>** |  |  | .24 |  |  | .24 |  |  | .26 |  |  | .37 |
| **F for change in R<sup>2</sup>** |  |  | 176.90*** |  |  | 1.71 |  |  | 4.60*** |  |  | 16.70*** |

Notes: PHQ-2 = Patient Health Questionnaire-2; SSCS-S = Self-Compassion Scale; SOC-3 = Sense of Coherence; ESSI = ENRICH Social Support Instrument.
All values are standardized regression weights. \* $p < .05$ ; \*\* $p < .01$ ; \*\*\* $p < .001$ .

In the first model, entering baseline depressive symptoms significantly explained 24% of variance (β=0.49, p<.001). Adding sociodemographic characteristics in Model 2 did not contribute to explain a significant added variance. Adding job demands in Model 3 produced an added 2% of explained variance, with risk perception (T2) scores showing greater β value of 0.13 (p=.005). Adding the individual resources in Model 4 produced an added 9% of explained variance, with SCS (T2), SOC (T2), ESSI (T1 and T2) scores showing greater β values of −0.24, −0.18, 0.11, −0.19 respectively (p<.001; p<.001; p=.04; p<.001, respectively). The final model predicted 37% of the explained PHQ-2 variance.

A hierarchical regression analysis was performed to assess which variables contribute to explain the anxiety symptoms at the T2 (see Table 5).

**Table 5.**
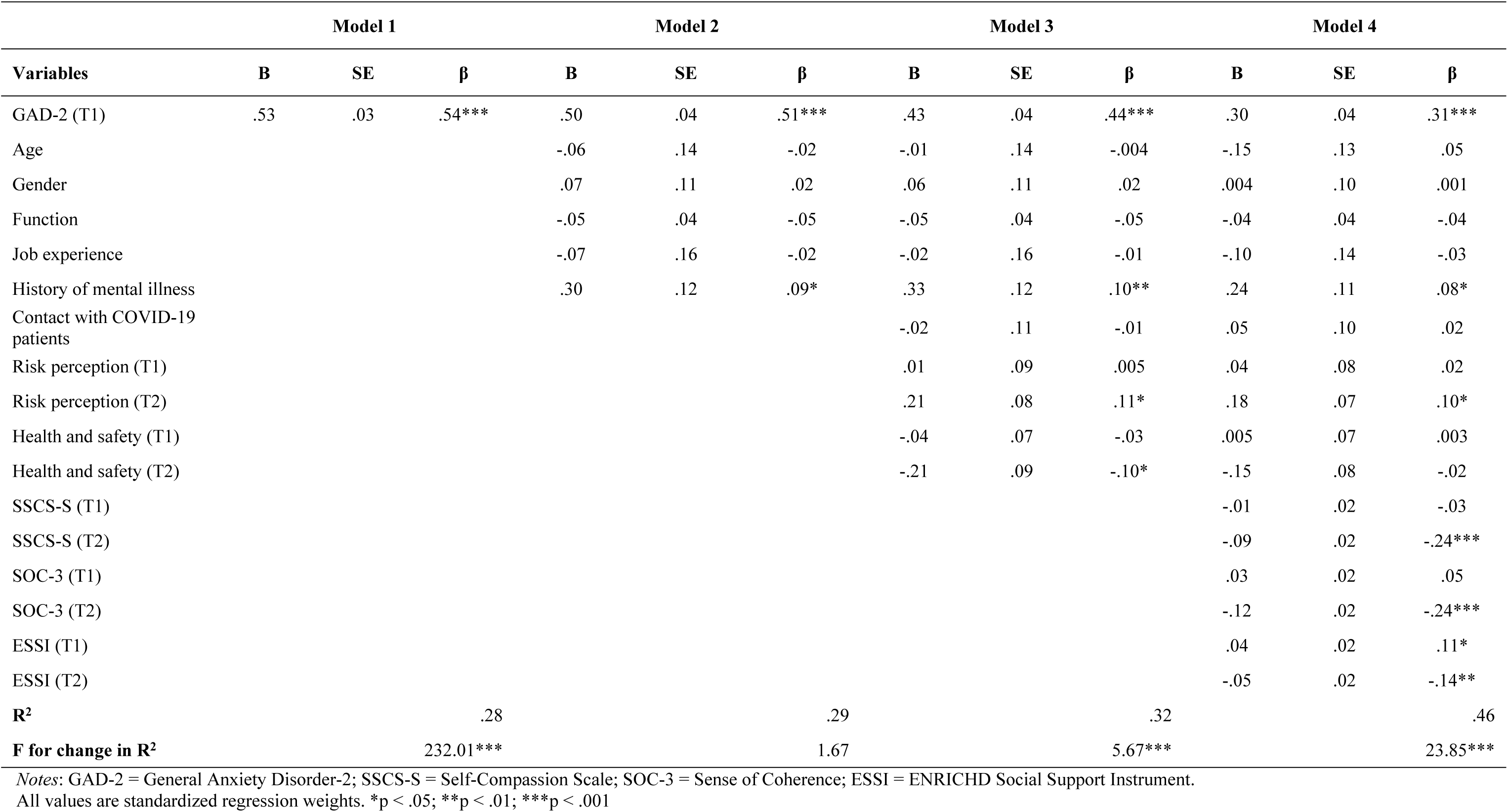
Summary of hierarchical regression model predicting anxiety symptoms (GAD-2) at T2.

|  | Model 1 |  |  | Model 2 |  |  | Model 3 |  |  | Model 4 |  |  |
| --- | --- | --- | --- | --- | --- | --- | --- | --- | --- | --- | --- | --- |
| Variables | B | SE | $\beta$ | B | SE | $\beta$ | B | SE | $\beta$ | B | SE | $\beta$ |
| GAD-2 (T1) | .53 | .03 | .54*** | .50 | .04 | .51*** | .43 | .04 | .44*** | .30 | .04 | .31*** |
| Age |  |  |  | -.06 | .14 | -.02 | -.01 | .14 | -.004 | -.15 | .13 | .05 |
| Gender |  |  |  | .07 | .11 | .02 | .06 | .11 | .02 | .004 | .10 | .001 |
| Function |  |  |  | -.05 | .04 | -.05 | -.05 | .04 | -.05 | -.04 | .04 | -.04 |
| Job experience |  |  |  | -.07 | .16 | -.02 | -.02 | .16 | -.01 | -.10 | .14 | -.03 |
| History of mental illness |  |  |  | .30 | .12 | .09* | .33 | .12 | .10** | .24 | .11 | .08* |
| Contact with COVID-19 patients |  |  |  |  |  |  | -.02 | .11 | -.01 | .05 | .10 | .02 |
| Risk perception (T1) |  |  |  |  |  |  | .01 | .09 | .005 | .04 | .08 | .02 |
| Risk perception (T2) |  |  |  |  |  |  | .21 | .08 | .11* | .18 | .07 | .10* |
| Health and safety (T1) |  |  |  |  |  |  | -.04 | .07 | -.03 | .005 | .07 | .003 |
| Health and safety (T2) |  |  |  |  |  |  | -.21 | .09 | -.10* | -.15 | .08 | -.02 |
| SSCS-S (T1) |  |  |  |  |  |  |  |  |  | -.01 | .02 | -.03 |
| SSCS-S (T2) |  |  |  |  |  |  |  |  |  | -.09 | .02 | -.24*** |
| SOC-3 (T1) |  |  |  |  |  |  |  |  |  | .03 | .02 | .05 |
| SOC-3 (T2) |  |  |  |  |  |  |  |  |  | -.12 | .02 | -.24*** |
| ESSI (T1) |  |  |  |  |  |  |  |  |  | .04 | .02 | .11* |
| ESSI (T2) |  |  |  |  |  |  |  |  |  | -.05 | .02 | -.14** |
| **R<sup>2</sup>** |  |  | .28 |  |  | .29 |  |  | .32 |  |  | .46 |
| **F for change in R<sup>2</sup>** |  |  | 232.01*** |  |  | 1.67 |  |  | 5.67*** |  |  | 23.85*** |

Notes: GAD-2 = General Anxiety Disorder-2; SSCS-S = Self-Compassion Scale; SOC-3 = Sense of Coherence; ESSI = ENRICH Social Support Instrument. All values are standardized regression weights. \*p < .05; \*\*p < .01; \*\*\*p < .001

In the first model, entering baseline anxiety symptoms significantly explained 28% of variance (β=0.54, p<.001). Adding sociodemographic characteristics in Model 2 produced an added 1% of explained variance, with history of mental illness scores showing greater β value of 0.09 (p=.01). Adding job demands in Model 3 produced an added 3% of explained variance, with risk perception (T2) and health and safety (T2) scores showing greater β values of 0.11 and −0.10 respectively (p=.01, p=.02, respectively). Adding individual resources in Model 4 produced an added of 14% of explained variance, with SCS (T2), SOC (T2), ESSI (T1,T2) scores showing greater β values of −0.24, −0.24, 0.11, −0.14 (p<.001, p<.001, p=.03, p=.003, respectively). The final model predicted 46% of the explained GAD-2 variance.

## Discussion

This longitudinal study aimed to examine long-term changes in job demands and individual resources associated with emotional distress symptoms in hospital staff during the COVID-19 pandemic. The results of our survey point out some relevant aspects.

Alarmingly, our data showed that 59.9% to 71.2% of HCWs reported anxiety or depression symptoms during the COVID-19 pandemic. This result is in line with the literature showing a rate of distress symptoms of up to 71.3% among HCWs during the COVID-19 outbreak [19]. Our first important finding was that hospital staff showed an overall increase in depressive (21.9%) and anxiety (17.8%) symptoms between the first and second waves of the COVID-19 pandemic. This finding suggests that the burden for the hospitals during the COVID-19 pandemic had a long-term impact on HCWs’ emotional distress. Previous longitudinal studies have shown that HCWs have experienced high levels of anxiety, depression, burnout, and post-traumatic stress disorder (PTSD) as a result of their work during the pandemic [42–44]. The prolonged nature of the pandemic, as well as long working hours, exposure to critically ill patients, and the risk of contracting the virus itself, added to the stress and emotional toll on HCWs [45].

In our first hypothesis, we expected that HCWs with higher levels of anxiety and depression over time would show higher levels of job demands and lower individual resources during both the first and second phases of the COVID-19 pandemic. This hypothesis was partially confirmed.

Compared with the other groups, individuals with high symptoms of depression or anxiety at both T1 and T2 were more likely to report a previous history of mental illness and lower levels of self-compassion and sense of coherence over time. Contrary to previous studies [e.g.,46], no differences were found between groups on levels of social support and job demands (i.e., contact with COVID-19 patients, risk perception, and health and safety). Noteworthily, the level of subjective stress due to lower psychological resources seems to be more related to emotional symptoms than the objective level of stress due to work-related factors. Recently, a study on 7,765 HCWs in Germany found that high levels of psychological resources were negatively associated with symptoms of anxiety and depression [47], even and above the effect of sociodemographic and work-related factors (such as female gender or contact with COVID-19- infected patients). There is evidence that psychological resources, such as self-compassion [48,49] and a sense of coherence [50–52], may be strongly related to lower distress symptoms in hospital staff. HCWs who practice self-compassion may be better able to accept difficult emotions and experiences and maintain perspective during the pandemic [48,49]. On the same line, HCWs who have a high sense of coherence seem to have greater coping abilities and resilience in the face of stress [53].

It is also interesting to note that anxiety, but not depressive, symptoms over time were less present within physicians and more experienced workers in our sample. It is possible to suppose that physicians and staff with greater professional experience have specific resilience to psychological pressure that might be related to professional fulfillment [54], self-empowerment [55] and greater experience with complex clinical problems and challenging situations.

Consistent with our second hypothesis, we found that risk perception, self-compassion, and sense of coherence at T2 and social support at both T1 and T2 were strong independent predictors of both depressive and anxiety symptoms at T2, even after controlling for baseline depressive or anxiety symptoms and sociodemographic variables. In addition, having a history of mental illness was a significant predictor of anxiety symptoms at T2. Once again, these results suggest that emotional symptoms are influenced more by psychological resources than by sociodemographic or work-related factors. Our finding is consistent with a growing number of studies that have investigated the role of psychological resources, such as resilience and coping strategies, as protective factors for HCWs during the COVID-19 pandemic [42,43]. A systematic review of 31 studies found that HCWs who reported higher levels of psychological resilience, optimism, and positive coping strategies, such as problem-solving and active coping, were less likely to experience symptoms of anxiety and depression during the pandemic [24]. Additionally, HCWs who reported higher levels of social support from colleagues and supervisors also had lower levels of anxiety and depression [56]. Another study found that HCWs who reported higher levels of emotional intelligence, self-compassion, and mindfulness were less likely to experience burnout and PTSD symptoms during the pandemic [57].

Although the study has the advantage of using a longitudinal design, some limitations are to be acknowledged. First, the number of participants who had responded to both T1 and T2 was relatively low compared to both cross-sectional surveys. This may be related to the characteristics of recruitment: HCWs were explicitly invited to participate in the survey at T2, even though they had not responded to the first questionnaire at T1. Second, our sample of HCWs included an unbalanced proportion between females and males and between the different countries. This surely limits the generalizability of the results. Third, the online administration of a survey is subject to responder bias. Particularly with HCWs during the dramatic pandemic, people who agreed to answer questions on their psychological health might be much more motivated to participate if psychologically distressed. This may have overestimated the emotional distress. Fourth, although most of the measures used have been validated, some questions were constructed ad hoc for the survey (e.g., job demands). Even though this may be a helpful way to measure, given time constraints, their validity in the current context may be up for debate. Finally, many possible confounding variables were not assessed and could not be controlled for, such as personality traits, PTSD symptoms or sleep disturbances.

Overall, this multicountry study pointed out that HCWs during the COVID-19 outbreak experienced a high burden of psychological distress. Regardless of personal vulnerability, relying on individual and contextual resources helps HCWs to cope with high-risk emergencies such as the COVID-19 pandemic. Our results strongly highlight that the mental health and resilience of HCWs should be supported during disease outbreaks by instituting workplace interventions for psychological support. Moreover, there is evidence that interventions focused on improving individual resources (such as mindfulness programs) helps HCWs cope with the health emergency situation [58].

## Data Availability

The data sets generated during and/or analysed during the current study are available from the corresponding author on reasonable request.

## Acknowledgments

Members of the Cope-Corona Study Group in alphabetical order: Baillès, Eva (Hospital Universitari Vall d’Hebron, Department of Mental Health, Barcelona, Catalonia, Spain and Hospital Nostra Senyora de Meritxell, Escaldes-Engordany, Andorra); Blanch, Jordi (Hospital Clínic de Barcelona, Servei de Psiquiatria i Psicologia, Barcelona, Spain); Cañizares, Silvia (Hospital Clínic de Barcelona, Servei de Psiquiatria i Psicologia, Barcelona, Spain); Cervera Teluel, Marta (Dexeus University Hospital, Department of Psychiatry Psychology and Psychosomatics, Barcelona, Spain); Dunne, Padraic J (Royal College of Surgeons in Ireland, University of Medicine and Health Sciences, Centre Of Positive Psychology and Health, Dublin, Ireland); Fadgyas Stanculete, Mihaela (University of Medicine and Pharmacy Iuliu Hatieganu, Cluj-Napoca, Romania); Farré, Josep Maria (Dexeus University Hospital, Department of Psychiatry Psychology and Psychosomatics, Barcelona, Spain); Font, Elena (Hospital Clínic de Barcelona, Servei de Psiquiatria i Psicologia, Barcelona, Spain); Forner Puntonet, Mireia (Hospital Universitari Vall d’Hebron, Department of Mental Health, Barcelona, Catalonia, Spain); Fritzsche, Kurt (Universitätsklinikum Freiburg, Klinik für Psychosomatische Medizin und Psychotherapie, Freiburg, Germany); Gayán, Elena (Dexeus University Hospital, Department of Psychiatry Psychology and Psychosomatics, Barcelona, Spain); Huang, Mingjin (Sichuan Mental Health Center, Mianyang, China); Ibañez Jimenez, Pol (Hospital Universitari Vall d’Hebron, Department of Mental Health, Barcelona, Catalonia, Spain); König, Sarah (Social and Organizational Psychology, Catholic University Eichstätt-Ingolstadt, Eichstätt, Germany); Kundinger, Nina (Social and Organizational Psychology, Catholic University Eichstätt-Ingolstadt, Eichstätt, Germany); Lobo, Antonio (Universidad de Zaragoza, Departamento de Medicina y Psiquiatría, Zaragoza, Spain); Nejatisafa, Ali-Akbar (Tehran University of Medical Sciences, Psychosomatic Research Center, Department of Psychiatry, Tehran, Iran); Obach, Amadeu (Hospital Clínic de Barcelona, Servei de Psiquiatria i Psicologia, Barcelona, Spain); Offiah, Gozie (Royal College of Surgeons in Ireland, University of Medicine and Health Sciences, Dublin, Ireland); Parramon, Gemma (Hospital Universitari Vall d’Hebron, Department of Mental Health, Barcelona, Catalonia, Spain); Peri, Josep Maria (Hospital Clínic de Barcelona, Servei de Psiquiatria i Psicologia, Barcelona, Spain); Ramos Quiroga, Josep Antoni (Hospital Universitari Vall d’Hebron, Department of Mental Health, Barcelona, Catalonia, Spain); Rousaud, Araceli (Hospital Clínic de Barcelona, Servei de Psiquiatria i Psicologia, Barcelona, Spain); Schuster, Sara (Social and Organizational Psychology, Catholic University Eichstätt-Ingolstadt, Eichstätt, Germany); Szcześniak, Dorota (Department of Psychiatry, Wroclaw Medical University, Wrocław, Poland); Torres Mata, Xavier (Hospital Clínic de Barcelona, Servei de Psiquiatria i Psicologia, Barcelona, Spain); Xiong, Nana (Peking University Sixth Hospital, Peking University Institute of Mental Health, NHC Key Laboratory of Mental Health (Peking University), National Clinical Research Center for Mental Disorder (Peking University Sixth Hospital), Beijing, China). These authors contributed equally to this work.

## Ethics Statement

All participants completed the survey anonymously and provided online informed consent to take part in the study. A self-generated identification code was used to match subjects at different assessment points in time. No IP addresses or geographic data was gathered. Participants were informed about privacy, ethical aspects and data treatment and they could cease the process at any time. The study was designed and carried out in accordance with the World Medical Association Declaration of Helsinki and its subsequent revisions [33] and approved by the Institutional Review Board of Paracelsus Medical University, General Hospital Nuremberg (IRB-2020-017).

## Notes

### Competing Interest Statement

The authors have declared no competing interest.

### Funding Statement

The authors received no specific funding for this work.

